# Oxygen Saturation or breathlessness? – how best to predict pneumonia in patients presenting with a cough to primary care

**DOI:** 10.1101/2020.09.09.20191650

**Authors:** Elspeth Chambers, Beth Stuart, Michael Moore, Mark Lown

## Abstract

**Background:** Distinguishing pneumonia from acute bronchitis has an important bearing on treatment decisions. The best validated diagnostic decision rule was derived from the GRACE-09 study. The 3Cs study provides oxygen saturations in addition to clinical signs and symptoms. Oxygen saturation is a predictor in hospital settings and may be beneficial in primary care.

**Objectives:** This project evaluates whether the data from GRACE-09 and 3Cs studies can be mapped onto each other and be combined dataset to refine the diagnostic accuracy of the GRACE-09 clinical prediction rule for the diagnosis of pneumonia.

**Patients and Methods:** GRACE-09 included 3106 patients with acute cough in a multicentre European study. 3Cs is a UK based study of 28883 patients with acute cough.

Binary logistic regression was used in the 3Cs dataset to determine which variables significantly predicted low oxygen saturation (<95%). Statistically significant predictors were determined using area under curve (AUC). This equation was used to model the probability of low oxygen saturation in the GRACE-09 dataset and subsequently included in the GRACE-09 model to determine whether low oxygen saturation is a comparable predictor to breathlessness in predicting pneumonia.

**Results:** For 3Cs, using all seven variables achieved an AUC for low oxygen saturation of 0.73 (95% CI 0.72-0.74). Substituting low oxygen saturation into GRACE-09 model gives AUC 0.75 (95% CI 0.69-0.82) compared to original GRACE-09 using breathlessness and gives AUC 0.77 (95% CI 0.73-0.82).

**Conclusions:** Low oxygen saturation and breathlessness are similarly predictive and could be used interchangeably in a modified GRACE-09 prediction model.

**KEY MESSAGES:** Can pulse oximetry replace subjective breathlessness assessment in the GRACE-09 prediction rule? Pulse oximetry gives an objective measurement that can be used to assist the diagnosis of pneumonia in primary care.

## INTRODUCTION

Acute lower respiratory tract infections (LRTI) are a major cause of preventable deaths worldwide.^1^ It has been found that the majority of patients who present to primary care with an acute cough or LRTI are diagnosed as having acute bronchitis and the minority as having pneumonia.^2–6^ Acute bronchitis is a self-limiting condition with limited benefit from antibiotics, whereas pneumonia is a more serious illness that requires treatment with antibiotics as, if left untreated, it can be fatal.^6–9^

The yearly prevalence of community acquired pneumonia (CAP) in adults in the UK is 0.3% in a primary care setting.^10–12^

Many of the symptoms of pneumonia are non-specific and shared with other conditions, such as bronchitis and asthma, consequently the clinical diagnosis of pneumonia is difficult and is likely to miss many radiographic diagnoses.^6, 13^ Whilst NICE suggests that chest x-ray (CXR) is considered to be the gold standard for the diagnosis of pneumonia this is not routinely available in primary care.^14^ Patients with pneumonia often present with raised temperature (≥37.8°C), crackles on auscultation, raised pulse (>100 beats/min) and reduced blood oxygen saturation (< 95%), but these may also be signs of acute bronchitis or other self-limiting illnesses.^6, 15, 16^ Clinically distinguishing between pneumonia and other conditions in primary care without further investigations is challenging. With this diagnostic uncertainty GPs opt to over-treat rather than undertreat.^13, 17–19^ In addition any short-term benefits of antibiotic treatments should be balanced against the risks of side-effects (21% of people reported moderate side effects and 5% major side effects) and possible increase in resistance due to overuse or misuse.^3, 20, 21^ A British primary care systematic review suggested that GPs prescribe an antibiotic if there is diagnostic uncertainty.^22^ The fear of missing an important diagnosis results in increased antibiotic prescribing, exacerbated by the absence of effective predictor algorithms.^6, 22^ Diagnostic uncertainty is one of the drivers of antibiotic overprescribing and hence improved precision in diagnostic algorithms may be helpful to counter this.^14^

Prediction rules can be used to guide clinical decision making about antibiotic prescribing.^23, 24^ Currently point of care tests are not widely used in the UK. Therefore, prediction rules which enable decision-making based on a combination of signs and symptoms rather than diagnostic tests, may be more useful.^18, 25^ Prediction rules are useful in increasing diagnostic accuracy for pneumonia and assist by excluding those patients at low risk of pneumonia, thus reducing inappropriate antibiotic prescribing.^13^ Two prediction rules currently available to aid in the diagnosis of pneumonia in primary care are Genomics to combat Resistance against Antibiotics in Community-acquired LRTI in Europe (GRACE-09) and Cough Complication Cohort (3Cs). These both combine physical signs and symptoms with blood test results.^6, 15, 16^

Previous paediatric studies have investigated the clinical signs predictive of low oxygen saturation. In children the best predictors of hypoxaemia, are accessory muscle use and pulsus paradoxus.^29^ Other signs predictive of low oxygen saturation are cyanosis (highly specific but low sensitivity) and increased respiratory rate of ≥60 breaths/min.^1, 29–31^ These clinical signs are different to those found to be predictive of hypoxaemia in adults so are of limited applicability in an adult population.

The 3Cs prediction rule includes pulse oximetry, now widely available in primary care but not available during the earlier GRACE-09 study. Instead, the GRACE-09 study included information on breathlessness. As recruitment was from routine consultations involving a wide range of doctors and practices, it is felt to be a large generalisable cohort study.^16^

In the time limited GP consultation, a prediction rule assisting in the identification of highest risk patients for pneumonia could improve antibiotic stewardship.^20, 32^ There are limitations to both of the existing prediction rules. A more complete dataset would include pulse oximetry and a CXR for every participant. Both the 3Cs and GRACE-09 studies are important investigations which have potential to significantly influence appropriate primary care prescribing for LRTIs.

This project seeks to evaluate whether the data from these two studies can be mapped onto each other and if the combined information can improve the accuracy of the diagnosis of pneumonia.

Our objective was to evaluate whether the data from GRACE-09 and 3Cs studies can be mapped onto each other and whether the combined dataset can be used to refine the diagnostic accuracy of the GRACE-09 clinical prediction rule for the diagnosis of pneumonia.

## PATIENT AND METHODS

### Study Design

This study sought to determine which variables, present at initial consultation, were best predictive of oxygen saturation in the 3Cs dataset and whether this modelled value of oxygen saturation could be used in the GRACE dataset to predict pneumonia.

### Participants - GRACE

A cohort of 3106 patients with acute cough were recruited from 294 GPs in 16 primary care research networks in 12 European countries during three winters between 2007 and 2010.^6^

The presenting features of the patient’s signs and symptoms, comorbidities, medication use and the working clinical diagnosis at presentation was documented. Patients had a CXR within 7 days, which was reported by a radiologist’s blind to the patient’s clinical information. The report was categorised as: “normal CXR”, “acute bronchitis”, “lobar or bronchopneumonia” or “other”. GPs were not informed of the diagnosis until after the study was completed unless the CXR found consolidation or any other diagnosis that required further investigation.^6^

### Inclusion Criteria

Patients were included if they were aged 18 and older, presented with an acute or worsened cough (≤28-day duration) that was considered to be caused by LRTI by the GP.^6^

### Exclusion Criteria

They were excluded if this was a second presentation for this same illness episode, if they were pregnant, breastfeeding or were immunocompromised.^6^

### Participants - 3Cs

A cohort of 28883 patients were recruited between 2009-13 from 522 practices in the UK that presented with an acute cough that was recognised as a LRTI. Of these patients 720 (2.4%) had a CXR within the first 7 days.^16^

3Cs is a prospective cohort study. The presenting features and management were documented by the physician. Data was collected on age, smoking history, prior duration of symptoms, nature and the severity of symptoms, examination as well as a rating of illness severity and antibiotic prescribing.^16, 21^

Medical records were reviewed to determine CXR results and additional data on re-consultations and hospitalisation or death during the 30 days after the initial consultation.^16^

### Inclusion Criteria

Patients were included if they were aged 16 and older who presented with a new illness. The main presenting symptom was a new or worsening cough for 3 weeks or less (acute cough), that was judged to be infective in origin by the physician.^9, 16^

### Exclusion Criteria

Patients were excluded if they presented to the physician with other causes of acute cough, such as heart failure; they were unable to complete a patient diary, due to severe mental illness or dementia; had presented previously with the same episode of illness; were immunocompromised.^16^

Characteristics in the derived decision rules from the two studies are shown in Table 1.

**Table 1.** Comparison of clinical findings predictive of pneumonia in GRACE-09 and 3Cs cohort studies^6, 15, 16^

| Clinical Findings | GRACE-09 | 3Cs |
| --- | --- | --- |
| Pulse ($>100$ beats/min) | ✓ | ✓ |
| Temperature ($\geq 37.8^{\circ}\text{C}$) | ✓ | ✓ |
| C-Reactive Protein (CRP) ($>30\text{mg/L}$) | ✓ | |
| Diminished breath sounds on auscultation | ✓ |  |
| Absence of runny nose | ✓ |  |
| Presence of breathlessness | ✓ |  |
| Crackles on auscultation | ✓ | ✓ |
| Oxygen saturation ($<95\%$) | | ✓ |

### Statistical Analysis

#### Step 1 – Prediction of oxygen saturation in the 3Cs study

Oxygen saturation was assessed in the 3Cs study using pulse oximetry. The distribution of oxygen saturations recorded in primary care is a highly skewed variable, with most values being 97% or higher (Figure 1) so it is inappropriate to use a linear model. Given the need for GPs to rule out those at low risk and rule in those at high risk for further investigation or treatment, it was appropriate to treat oxygen saturation as a binary variable. For the purposes of this study, impaired oxygen saturation was defined as a value of less than 95% (as in the 3C study).^16^ Potential predictors included patient characteristics (age, gender and medical history), patient reported symptoms at presentation and clinical signs found on examination of the patient at the primary consultation.

**Figure 1.**
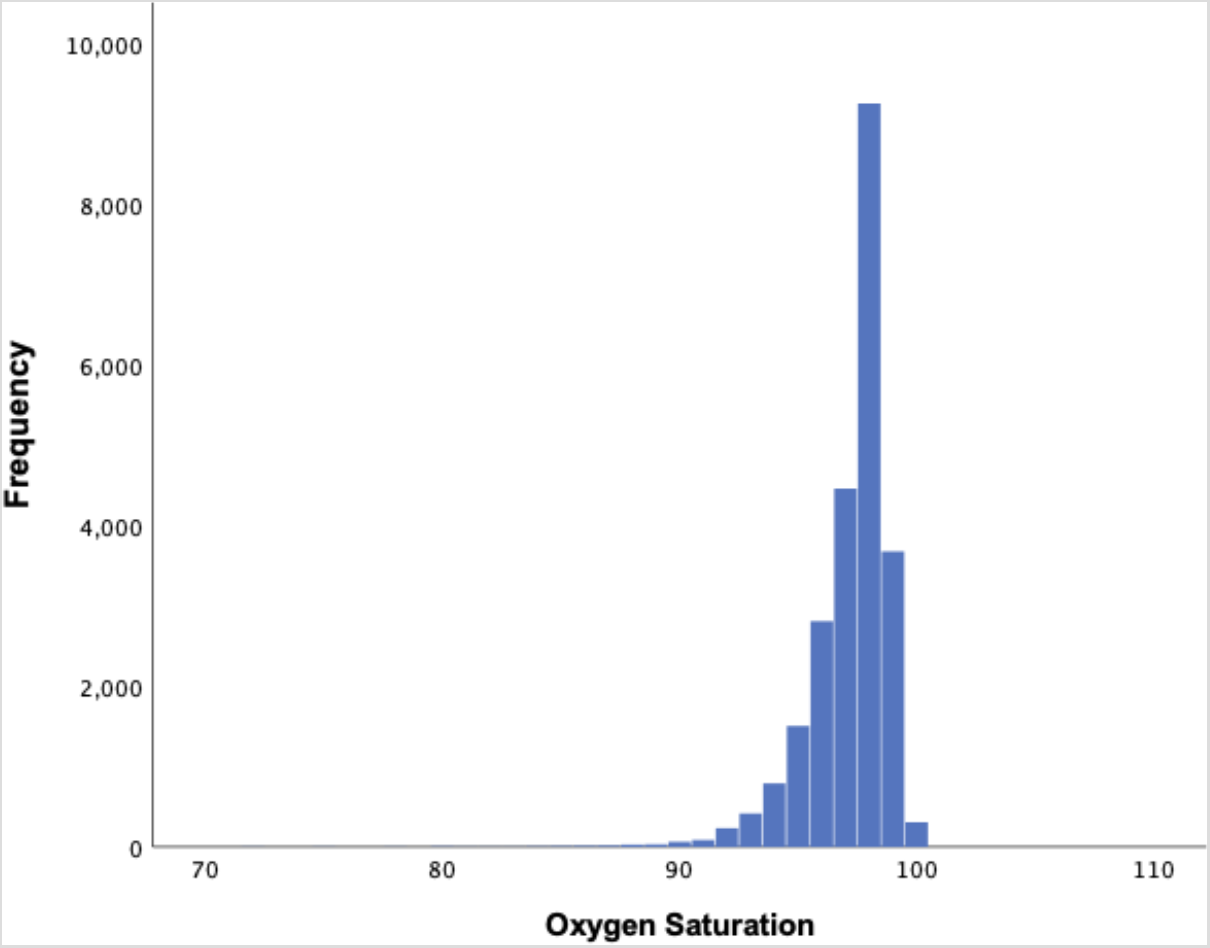
Histogram showing distribution of oxygen saturation in the 3Cs dataset

To determine which variables were statistically significant in the prediction of low oxygen saturation, a binary logistic regression model was used. The predictive value of statistically significant predictors from the logistic regression model was then determined using area under the receiver operating curve (AUC). Variables were sequentially added to the receiver operating characteristic (ROC) curve analysis starting with the most predictive in a univariate analysis.

#### Step 2 – Prediction of oxygen saturation in the GRACE study

The regression equation from the model developed from the 3Cs study was then applied to the participant data in the GRACE dataset to model probability of low oxygen saturation. The output of logistic regression is the predicted probability of low oxygen saturation.

To determine the optimal cut-point for low oxygen saturation the Liu method was used to select the point that maximised the sensitivity and specificity.^33^

This imputed probability of low oxygen saturation was then included in the prediction rule and the AUC analysed to determine whether it improved the ability of the GRACE-09 prediction model to rule in and rule out pneumonia.

The calibration of both the models was assessed using the Hosmer-Lemeshow statistic.^34^

## RESULTS

### Participant Characteristics

Both the 3Cs and GRACE datasets are from existing completed prospective cohort studies. The key characteristics of the study populations are summarised below in Table 2. The patient characteristics in both datasets are similar. In both there was an over representation of female participants. The majority in both datasets were below 60 years old.

**Table 2.** Baseline characteristics of study participants in 3Cs and GRACE cohort studies

|  | 3Cs | GRACE |
| --- | --- | --- |
| **Age ($\geq 60$ years)** | 10886/28846 (37.7%) | 992/3112 (31.9%) |
| **Female** | 17098/28841 (59.3%) | 1512/2533 (59.7%) |
| **Any comorbidity** | 13100/28846 (45.4%) | 859/3104 (27.7%) |
| **Lung comorbidity** | 7461/28846 (25.9%) | 531/3107 (17.1%) |
| **Never smoked** | 13212/28377 (46.6%) | 1078/1754 (61.5%) |
| **Influenza Vaccination** | 9842/28846 (34.1%) | 735/3107 (23.7%) |
| **Symptoms** |  |  |
| Breathlessness | 19665/28844 (68.2%) | 1756/3106 (56.5%) |
| Fever | 15085/28845 (52.3%) | 1087/3104 (35.0%) |
| Cough | 28808/28841 (99.9%) | 3103/3106 (99.9%) |
| Chest pain | 11777/28845 (40.8%) | 1437/3104 (46.3%) |
| Headache | 16477/28845 (57.1%) | 1744/3107 (56.1%) |
| Muscle ache | 13585/28844 (47.1%) | 1574/3106 (50.7%) |
| Sleep disturbances | 23481/28844 (81.4%) | 1906/3105 (63.1%) |
| Confusion | 2313/28842 (8.0%) | 137/3106 (4.4%) |
| Diarrhoea | 3800/28845 (13.2%) | 222/3106 (7.1%) |
| Sputum | 21407/28845 (74.2%) | 2478/3105 (79.8%) |
| **Clinical Examination** |  |  |
| Respiratory rate ($\geq 30$ breaths/min) | 551/28846 (1.9%) | 23/3032 (0.8%) |
| Temperature ($\geq 37.8^\circ\text{C}$) | 1656/28825 (5.7%) | 169/3079 (5.5%) |
| Pulse ($> 100$ beats/min) | 2150/28834 (7.5%) | 86/3064 (2.8%) |
| Blood Pressure | 2193/28846 (7.6%) | 10/3041 (2.6%) |
| *Systolic BP ($< 90$ mmHg)* | 243/28846 (0.8%) | 79/3041 (0.7%) |
| *Diastolic BP ($< 60$ mmHg)* | 2122/28846 (7.4%) | 67/3041 (2.2%) |
| Wheeze | 7072/28837 (24.5%) | 1329/3105 (42.8%) |
| Crackles | 12257/28839 (42.5%) | 290/3091 (9.4%) |

Overall, whilst there was a higher prevalence of some signs and symptoms in 3Cs than in GRACE-09, given that the inclusion and exclusion criteria are broadly similar it seems unlikely that the populations examined are significantly different.

### Predictors of low oxygen saturation in the 3Cs dataset

Logistic regression analysis of 3Cs data suggested that the significant predictors of a low oxygen saturation were “ever smoked”, wheeze, lung comorbidity (asthma, COPD or other lung disease), confusion, gender, respiratory rate and age (Table 3).

**Table 3.** Variables predictive of low oxygen saturation in 3Cs dataset

| Predictive Variables | B | 95% Confidence Interval |  |
| --- | --- | --- | --- |
|  |  | Lower | Upper |
| **Ever smoked** | -0.448 | 0.572 | 0.713 |
| **Wheeze** | -0.714 | 0.440 | 0.546 |
| **Lung comorbidity** | -0.424 | 0.586 | 0.730 |
| **Confusion** | -0.259 | 0.646 | 0.922 |
| **Gender** | -0.213 | 0.728 | 0.896 |
| Age | 0.032 | 1.029 | 1.036 |
| Respiratory rate | 0.045 | 1.035 | 1.057 |
| Constant | -1.789 |  |  |

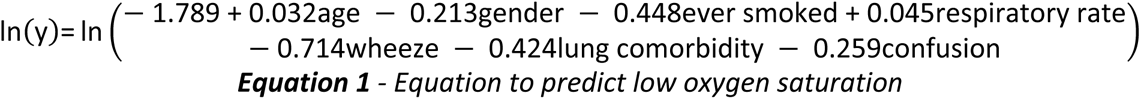

The variables were added sequentially into an AUC curve analysis, beginning with the most predictive and the AUC is presented for each step in Table 4.

**Table 4.** AUC of successive statistical models combining the significantly predictive clinical signs from 3Cs dataset

| Predictive Variables | AUC (95% CI) |
| --- | --- |
| Age | 0.67 (0.65-0.68) |
| Age + Gender | 0.67 (0.67-0.68) |
| Age + Gender + Ever Smoked | 0.69 (0.67-0.70) |
| Age + Gender + Ever Smoked + Respiratory Rate | 0.70 (0.70-0.71) |
| Age + Gender + Ever Smoked + Respiratory Rate + Wheeze | 0.73 (0.71-0.74) |
| Age + Gender + Ever Smoked + Respiratory Rate + Wheeze + Lung Comorbidity | 0.73 (0.72-0.74) |
| Age + Gender + Ever Smoked + Respiratory Rate + Wheeze + Lung Comorbidity + Confusion | 0.73 (0.72-0.74) |

The final model including all seven variables gave an AUC of 0.73 (95% CI 0.72-0.74) (Figure 2). The Hosmer-Lemeshow test value of p = 0.581 indicates that this data is well calibrated for determining low oxygen saturation.

**Figure 2.**
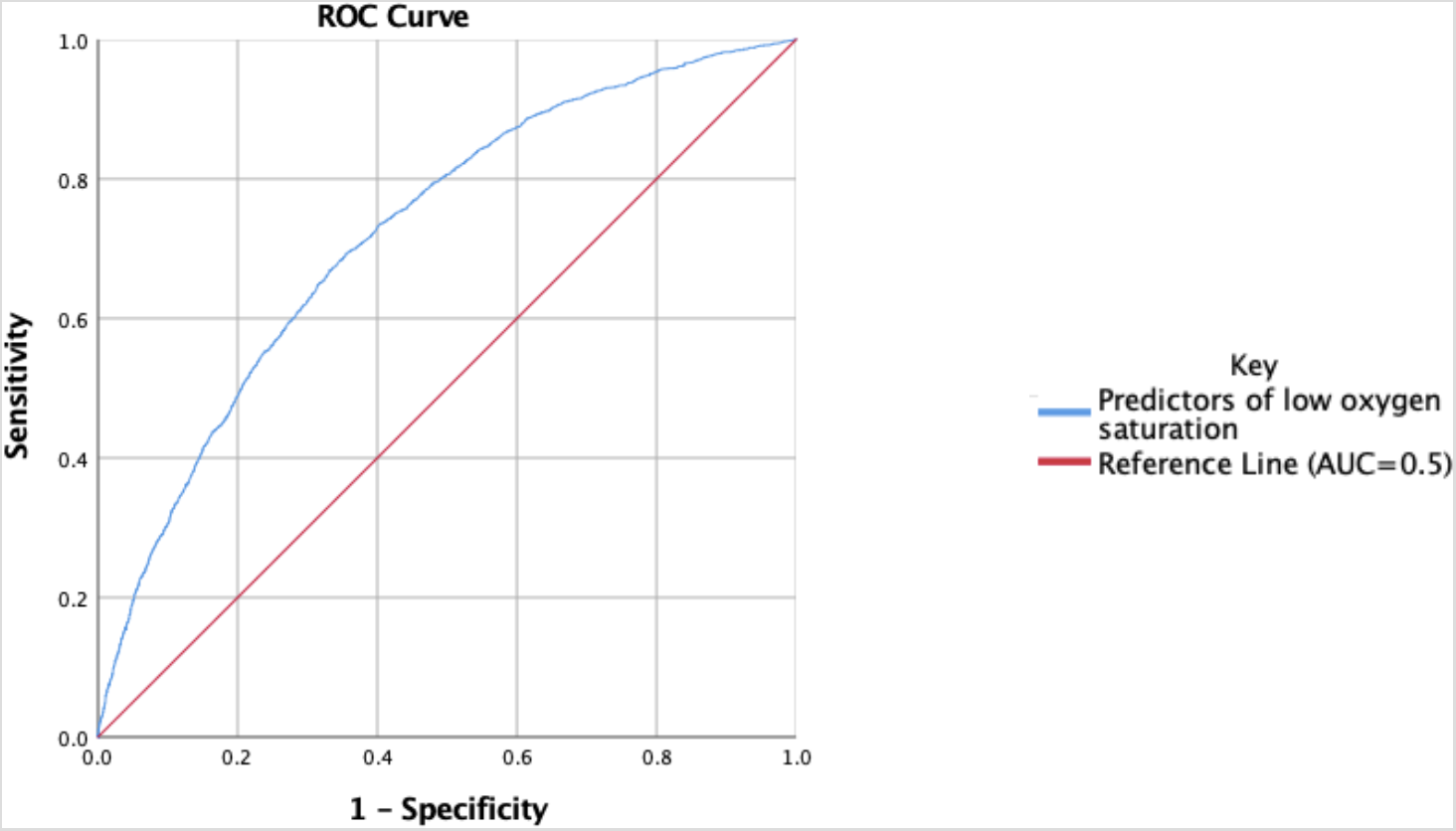
ROC curve showing AUC of variables predictive of low oxygen saturation in 3Cs dataset.

The equation for predicting a low oxygen saturation that was generated by the AUC analysis (Table 3) and is shown above in Equation 1.

### Predicting low oxygen saturation in the GRACE dataset

Using the equation derived from the 3Cs dataset above (Equation 1) enabled prediction of the probability of low oxygen saturations in the GRACE dataset. With this modelled value of low oxygen saturation, the GRACE-09 prediction rule was re-run (raised pulse (>100 bpm), raised temperature (≥37.8°C), raised CRP (>30mg/L), diminished breath sounds on auscultation, absence of runny nose, breathlessness, crackles on auscultation), but substituting low oxygen saturation in the revised rule for breathlessness in the original GRACE rule. The revised rule generates an AUC of 0.76 (95% CI 0.69-0.82) (Figure 4).

**Figure 3.**
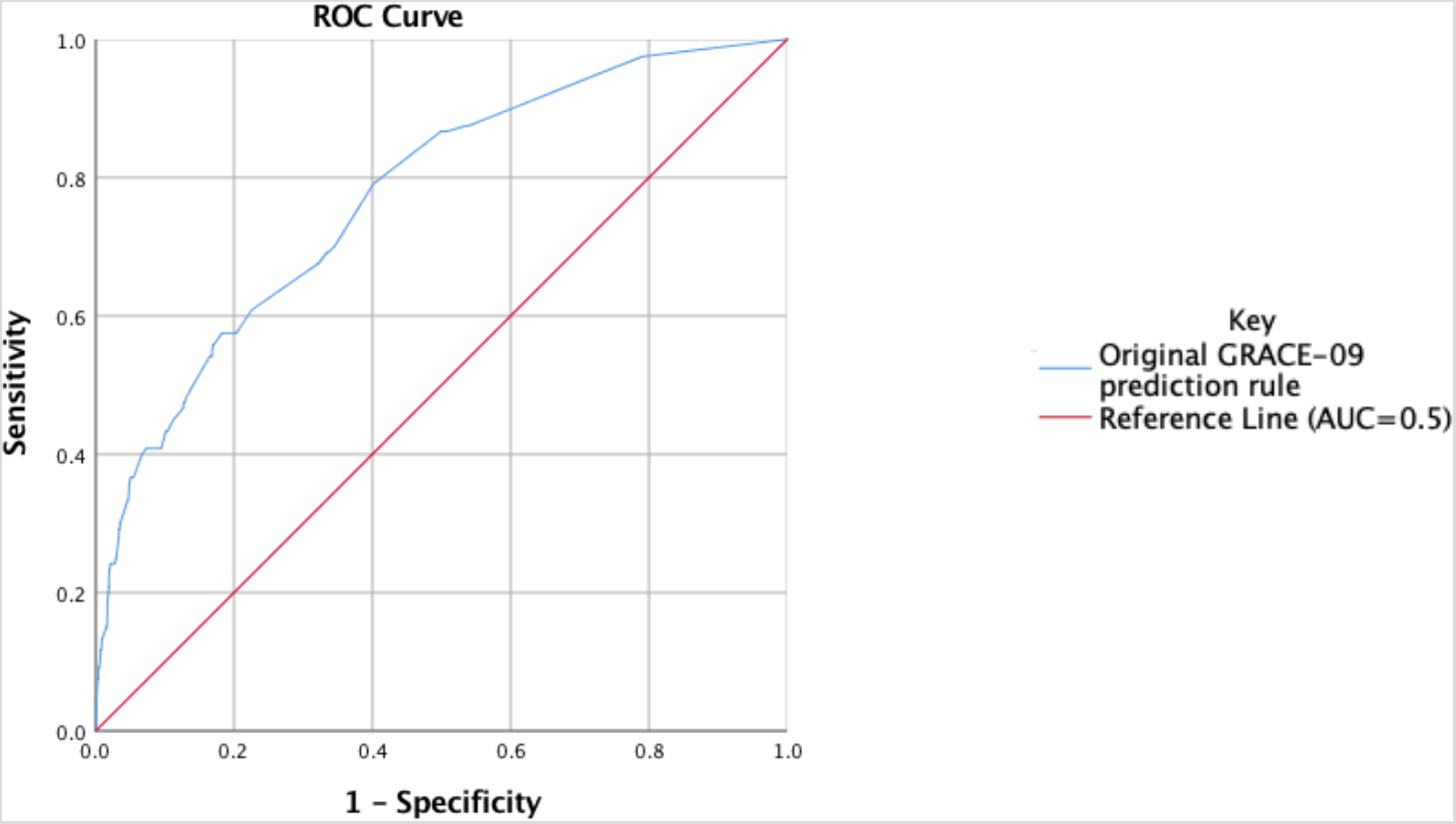
AUC using GRACE-09 prediction rule with breathlessness.

**Figure 4.**
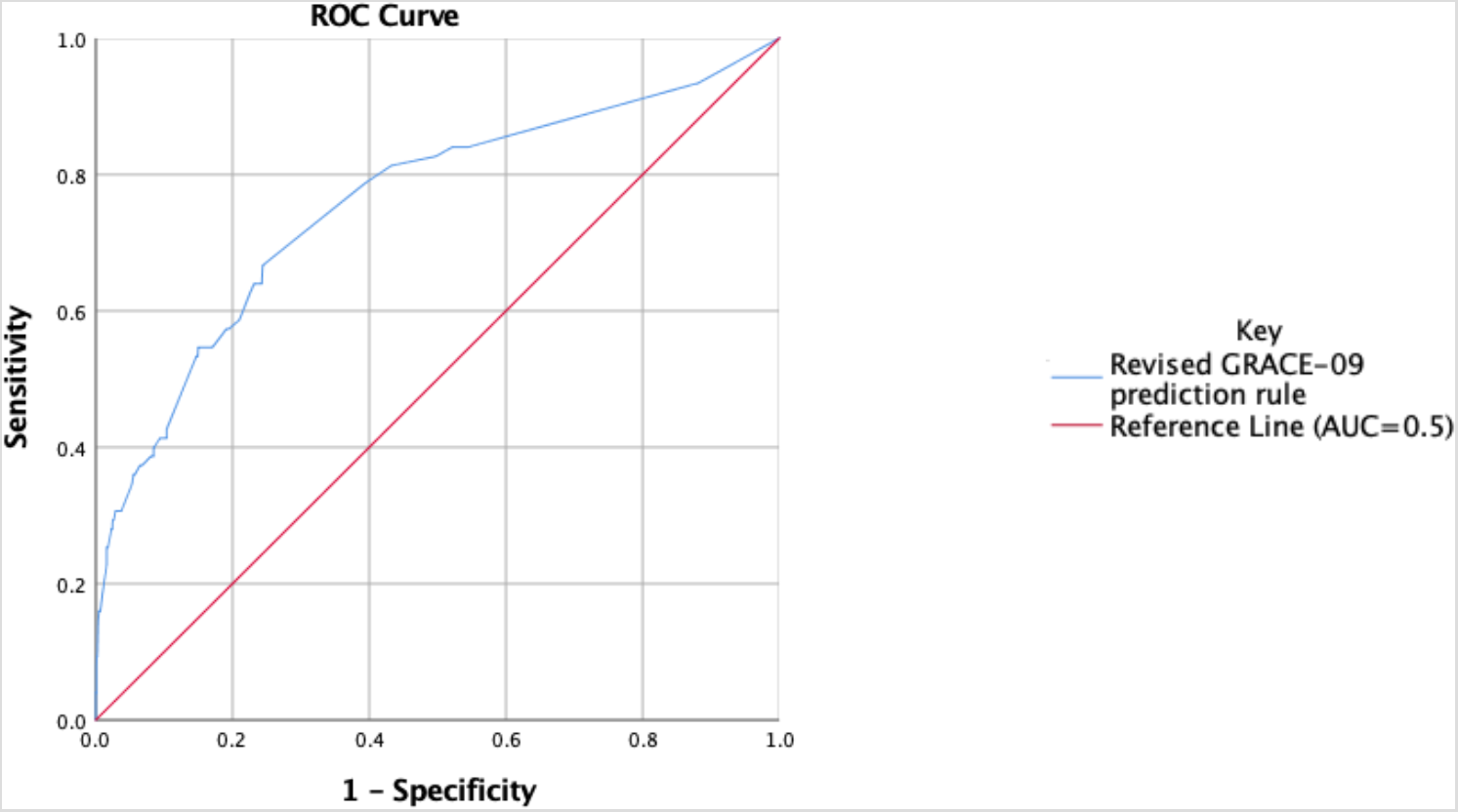
AUC using revised GRACE-09 prediction rule with binary value of low oxygen saturation

The revised rule replacing breathlessness with oxygen saturation can be compared to the original published GRACE-09 prediction rule which gave an AUC of 0.76 (95% CI 0.71-0.80) (Figure 3). The confidence intervals for these two rules are similar, suggesting there is no statistically significant difference between them.

The Hosmer-Lemeshow test value of p = 0.394 indicates that this revised GRACE-09 prediction rule (using a binary value for oxygen saturation) is well calibrated.

The original GRACE-09 prediction rule included measurement of CRP which is not routinely measured in a primary care setting in the UK.^35^ Therefore, further modelling using the GRACE dataset, but omitting the CRP test was undertaken and the AUC reduced to 0.71 (95% CI 0.66-0.76). Similar manipulation of the revised GRACE-09 prediction rule decreases the AUC to 0.70 (95% CI 0.63-0.77).

## DISCUSSION

### Main Findings

The research suggests that it is possible to map low oxygen saturation onto a combination of signs and symptoms, including age, gender, presence of wheeze, increased respiratory rate, history of smoking, lung comorbidity (asthma, COPD or other lung disease) and presence of confusion.

This study (using the GRACE dataset) concludes that when using a prediction rule for the diagnosis of community acquired pneumonia substituting low oxygen saturation for breathlessness results in a decision rule with similar performance to the original rule.

### Strengths and Limitations

Both the 3Cs and the GRACE are large cohort studies with large datasets including a significant number of signs, symptoms and clinical findings. Both studies recruited patients presenting in primary care, but it is impossible to exclude some selection bias due to the process of recruitment of participants.

The adult study population mean that findings should not be generalised to the paediatric population without further study. Whilst removing CRP from of the GRACE-09 prediction rule reduced the predictive value, it more accurately represents current practice in the UK.^18^ Similarly, treating low oxygen saturation as a binary variable means that this is an imputed value rather than an actual measured value. Further studies are required to confirm whether using measured values of oxygen saturation would give similar results. Ideally, such studies would need to collect data correlating oxygen saturations, breathlessness and CXRs. Some variables are missing for participants in both the GRACE and 3Cs datasets, potentially introducing bias, however when values were imputed in the original 3Cs study, this made no significant difference to the results generated.^16^ Whilst an AUC above 0.9 would be considered outstanding, the value of 0.76 is considered to be good.^34^.

### Overall Implications

There is currently poor correlation between the clinical and radiographic diagnosis of pneumonia. Whilst breathlessness is an important symptom for the patient to report, it is a subjective assessment and lacks objective correlation.^36^ The pulse oximeter however, assuming it is appropriately calibrated and accurate, will give a specific value which is indisputable. Pulse oximetry is an easily measured and robust technology. It is widely available in UK primary care and has few limitations (severe anaemia, hypovolemia, arrhythmias).^16, 37–39^

### Clinical Implications

A rule which is recognised as being highly predictive and easily utilised would assist in the accurate diagnosis and appropriate treatment of community acquired pneumonia. Diagnostic accuracy can be improved using a prediction rule based on a combination of the clinical signs and symptoms. Increasing the use of objective measurable variables such as oxygen saturation and reducing reliance on subjective symptoms increases the value of this predictive rule.^16^ This is relevant as an increasing number of clinical assessments are being performed by less experienced clinicians.

Antibiotic prescribing in patients presenting with cough to primary care should be limited to those with high risk of a diagnosis of community acquired pneumonia. Limiting the use of antibiotics to those at highest risk of pneumonia would lead to a reduction in the utilisation of antibiotics and reduce the threat of developing antimicrobial resistance.^19^

## Conclusion

Modification of an existing clinical decision rule to incorporate oxygen saturation in place of breathlessness has similar test performance. It is likely that breathlessness and oxygen saturation can be used interchangeably in the GRACE-09 prediction rules (original and revised) depending on the clinical situation.

## Data Availability

all data referred to in the manuscript is available

## ACKNOWLEDGEMENTS

This project was undertaken as a research project at University of Southampton Medical School and would not have been possible without the support, guidance and advice of all of my supervisors, Dr Beth Stuart, Dr Mark Lown and Professor Michael Moore.

## FUNDING

This was an unfunded student project.

## TRANSPARENCY DECLARATION

None to declare

## Notes

### Competing Interest Statement

The authors have declared no competing interest.

### Funding Statement

This was an unfunded Medical Student project

### Author Declarations

this was an unfunded medical student project with ethics approval from the university of Southampton

